# Valve Sparing Aortic Root Replacement: long term predictors of outcomes and mortality

**DOI:** 10.1101/2023.05.10.23289816

**Authors:** Varun J. Sharma, Abbie Kangarajah, Amy Yang, Michelle Kim, Siven Seevayanagam, George Matalanis

## Abstract

**Introduction:** In aortic root surgery, Valve Sparing Aortic Root Replacement (VSARR) is an attractive alternative by mitigating the risks inherent to prosthetic valves, however little is known about the variables that impact its performance and durability. We have reviewed our mid to long-term outcomes following VSARR and describe factors that impact patient survival and aortic valve reintervention and insufficiency.

**Methods:** A retrospective review of 284 consecutive patients undergoing VSARR between November 1999 and January 2022 at Austin Health, Melbourne, Australia, was undertaken, with median follow up of 6.43 ± 4.83 years, but up to 22.0 years.

**Results:** The median age at intervention was 60.0 years (IQR 48.0-67.0), of which 68 (23.9%) had bicuspid aortic valve (BAV) disease, 27 (9.5%) Marfan’s disease, 119 (41.9%) severe aortic root dilation (>50mm), and 155 (54.6%) severe aortic insufficiency at the time of intervention. The 30-day mortality was 1.8%, with freedom from mortality of 96.0% (95% CI 92.6-97.8%) at 5 years, 88.2% (95% CI 81.4-92.6%) at 10 years and 78.6% (66.2-86.9%) at 15 years. Freedom from aortic re-intervention was 92.2% (95% CI 87.7-95.2%) at 5 years, 79.8% (95% CI 71.8-85.8%) at 10 years, and 74.1% (63.5-82.0%) at 15 years. Predictors of re-intervention were concomitant leaflet repair (HR 8.13, 95%CI 1.07-61.7) and bicuspid valvulopathy (HR 2.23, 95% CI 1.07-4.68). The freedom from aortic insufficiency was 89.1% (95% CI 83.5-92.9%), 84.9% (95% CI 77.8-89.9%) and 80.7% (71.0-87.4%) at 5-, 10- and 15-years respectively.

**Conclusion:** VSARR has excellent long-term outcomes, with low mortality and re-intervention rates. Concomitant Leaflet repair and Bicuspid Aortopathy are the only long-term predictors of re-intervention.

## Introduction

Aortic root replacement (ARR) is the gold standard for the treatment of patients with aortic root disease but is associated with lifelong risks inherent to prosthetic valves. Mechanical prostheses carry the risks of lifelong anticoagulation and systemic thrombo-embolism, whilst bioprosthetic valves are limited by long-term structural deterioration and need for reoperation.

As an alternative, Valve sparing aortic root replacement (VSARR) mitigates these risks and is effective in simultaneously treating the proximal aneurysmal disease. There are increasing data supporting it as a viable alternative for aneurysmal disease, including those with genetic disorders such as Marfan’s Disease, and is the treatment of choice for young patients with aortic root aneurysms with normal or near normal aortic cusps [1, 2].

Long term data for VSARR is still scarce [2], with concerns regarding the durability of the spared aortic valve and risk of re-intervention. Factors that predict poor outcomes with VSARR are poorly delineated providing challenges in case selection, especially in patients with bicuspid or genetic aortopathies, severe (>50mm) root dilation, aortic insufficiency, acute type A aortic dissection (ATAAD) presentation, and those needing concomitant leaflet repair.

We retrospectively assessed our experience to assess immediate and long-term outcomes following valve-sparing aortic-root replacement and describe factors that predict insufficiency or re-intervention.

## Methods

### Ethical Statement

The study was approved by the Health and Research Ethics Committees (HREC) of Austin Health, Melbourne, Victoria to meet ethical and legal requirements, and individual consent was waived (HREC LNR/17/Austin/82).

### Study Design

This series is the institute’s 22-year experience with valve-sparing aortic root replacement. It represents a retrospective cohort of consecutive patients from November 1999 to January 2022 (Austin Health and Warringal Hospital, Heidelberg, Melbourne, Australia). Data from these interventions is entered in the ANZSCTS (Australia New Zealand Society of Cardiothoracic Surgeons) and hospital database. The ANZSCTS database is a bi-national database and registry provider, adopted for collecting cardiac surgical data in Australasia, which includes sections that anaesthetists, surgeons and perfusionists consecutively fill. Data is periodically checked for accuracy by independent database managers. At our institution it has a 100% rate of completeness. Patient data including demographic, co-morbidities, surgical details, and follow-up correspondence (until last known follow up) was collected from these databases with patient identifiers removed.

### Patient Population

All patients who underwent valve sparing aortic root replacement were included and stratified based on their underlying morphology, falling into bicuspid (BAV) or tricuspid (TAV) valve. Determination of morphology was based on pre-operative echocardiography and confirmed intra-operatively. It includes both emergent and elective priorities, and includes patients from both elective aneurysm repair, acute type A aortic dissection (ATAAD) and aneurysm rupture. All patients underwent routine 6-24 monthly clinical, and imaging follow up appointments, the notes of which were reviewed for outcome analysis including mortality. All surviving patients had review in the last two years.

### Study Outcomes

The study assessed freedom from all-cause mortality, with secondary outcomes assessing freedom from aortic re-intervention or severe insufficiency. All-cause mortality was defined as mortality from any cause, including all aortic, cardiac and non-cardiac causes. Aortic reintervention included all endovascular and open procedures to any portion of the aorta or aortic valve, including for aortic stenosis and regurgitation. Valve insufficiency was defined using ACC/AHA guidelines [3], where 0 was defined as absent, 1 as trivial, 2 as mild, 3 as moderate and 4 as severe. Insufficiency of 3 or greater was clinically significant.

The ANZSCTS database data manual [4] was used to define all pre- and peri-operative variables and events. Other data collected included valve morphology (BAV or TAV), genetic aortopathies including Marfan’s, severe aortic root dilation (>50mm at time of intervention), concomitant cardiac intervention, leaflet repair and severe aortic insufficiency at time of presentation.

### Statistical methods

The distribution of patient demographics, procedures and outcomes were assessed using count proportions or median averages with corresponding interquartile ranges. Kaplan Meier analysis was used to assess long term outcomes of mortality and re-intervention. A multivariable cox-regression analysis with forward stepwise elimination was used to identify predictors of mortality, re-intervention and aortic insufficiency, with a p value of less than 0.20 required for consideration and 0.05 required for retention in the final model. Only factors significant (p<0.05) from multivariable analysis are listed, and factors of ATAAD, Bicuspid Aortic Valve (BAV) disease, Marfan’s, Severe Root (>=50mm), Concomitant Aortic Procedure, Leaflet Repair and Severe Aortic Insufficiency were tested independently in the final model. A Fine-Gray model was used for competing risk analysis. All data was collected on MS Excel (Microsoft Corporation, 2010), and analysis was undertaken on Stata v15.0 (Statacorp, Texas, USA).

## Results

### Demographics

From November 1999 to January 2022, there were 284 consecutive patients who underwent valve-sparing aortic root replacement at our institution as described in Table 1. The median follow up was 6.43 ± 4.83 years, with up to 22.0 years follow up. The median age at intervention was 60.0 years (IQR 48.0-67.0). There were 68 patients (23.9%) with bicuspid aortic valve (BAV) disease, 27 (9.5%) patients with Marfan’s disease, 119 (41.9%) with severe aortic root dilation (>50mm), and 155 (54.6%) had severe aortic insufficiency at the time of intervention. Patients with BAV underwent intervention at a younger age (51.5 vs. 62.0 years) with smaller aortic root sizes (46.0 vs. 49.0 mm), with no difference in the proportion of BAV patients undergoing VSARR between the pre-and post-2010 eras.

**Table 1.** Demographics of all patients who underwent Valve-sparing aortic root replacement from November 1999 to January 2022. *Abbreviations: AI = Aortic Insufficiency, ATAAD = Acute Type A Aortic Dissection, BAV = Bicuspid Aortic Valve, EF = Ejection Fraction, NYHA = New York Heart Association, TAV = Tricuspid Valve*

|  | Total Patients<br>(n=284) | TAV<br>(n=216) | BAV<br>(n=68) | P<br>Value |
| --- | --- | --- | --- | --- |
| **Demographic** |  |  |  |  |
| Age (IQR) | 60.0 (48.0-67.0) | 62.0 (51.5-68) | 51.5 (40.0-61.0) | <0.01 |
| Emergent | 15 (5.3%) | 14 (6.5%) | 1 (1.5%) | 0.11 |
| Diabetes Mellitus | 14 (4.9%) | 11 (5.1%) | 3 (4.4 %) | 0.82 |
| Smoking | 89 (31.3%) | 72 (33.3%) | 17 (25.0%) | 0.20 |
| Hypertension | 142 (50.0%) | 115 (53.2%) | 27 (39.7%) | 0.05 |
| Cerebrovascular Disease | 11 (3.9%) | 9 (4.2%) | 2 (2.9%) | 0.65 |
| Ischaemic Heart Disease | 38 (13.4%) | 31 (14.4%) | 7 (10.3%) | 0.39 |
| **Cardiac History** |  |  |  |  |
| **NYHA Class** |  |  |  |  |
| I | 168 (59.2%) | 124 (57.4%) | 44 (64.7%) | 0.29 |
| II | 82 (28.9%) | 61 (28.2%) | 21 (30.9%) | 0.68 |
| III | 32 (11.3%) | 29 (13.4%) | 3 (4.4%) | 0.04 |
| IV | 2 (0.70%) | 2 (0.93%) | 0 (0.0%) | 0.43 |
| **Aortic Diameters (mm)** |  |  |  |  |
| Annulus | 29.0 (25.0-31.0) | 28.0 (24.0-30.0) | 31.0 (28.0-33.0) | <0.01 |
| Aortic Root | 48.0 (42.0-51.0) | 49.0 (43.0-52.0) | 46.0 (41.0-50.0) | 0.03 |
| Sinotubular Junction | 40.0 (36.0-45.0) | 40.0 (35.0-46.0) | 39.0 (37.0-42.0) | 0.69 |
| Ascending Aorta | 49.0 (41.0-54.0) | 49.0 (41.0-55.0) | 49.0 (44.0-52.0) | 0.94 |
| Maximal Size | 50.0 (47.0-55.0) | 51.0 (48.0-55.0) | 50.0 (44.0-52.0) | <0.01 |
| **Valvular Calcification** | 40 (14.1%) | 26 (12.0%) | 14 (20.6%) | 0.11 |
| **ATAAD** | 17 (6.0%) | 17 (7.9%) | 0 (0.0%) | 0.02 |
| **Marfans** | 27 (9.5%) | 25 (11.6%) | 2 (3.0%) | 0.03 |
| **Severe Root (>50mm)** | 119 (41.9%) | 95 (44.0%) | 24 (35.3%) | 0.21 |
| **Severe AI** | 155 (54.6%) | 124 (56.9%) | 32 (47.1%) | 0.15 |
| **Redo Intervention** | 10 (3.6%) | 9 (4.2%) | 1 (1.6%) | 0.32 |
| **Pre-operative EF** | 60 (59.0-60.0) | 60.0 (60.0-60.0) | 60.0 (56.0-60.0) | 0.90 |
| **Era of Intervention** |  |  |  |  |
| Pre 2010 | 86 (30.3%) | 70 (32.4%) | 16 (23.5%) | 0.17 |
| Post 2010 | 198 (69.7%) | 146 (68.2%) | 52 (76.5%) | 0.17 |

### Operative Characteristics

Of 284 valve sparing root replacements, the 95.1% (n=270) were re-implantation (David’s) procedures, with the remaining 4.9% (n=14) being remodelling (Yacoub) procedures (Table 2). Concomitant arch repair was undertaken in 20.4% (n=57) of the cohort. BAV patients were less likely to undergo a concomitant cardiac procedure (26.5% vs. 44.0%), but there were no significant differences in the type of valve sparing root replacement procedure (re-implantation or re-modelling) they underwent. Leaflet repair was required in 70.4% of patients, with a larger proportion of BAV (82.4%) requiring repair compared to TAV patients (66.7%). Repair was undertaken either to the free edge (with or without re-enforcement), commissure, or intervention via a wedge resection or midpoint repair (Table 2). There were no other significant differences in the operative characteristics of both BAV and TAV cohorts.

**Table 2.** Operative characteristics of patients undergoing Valve Sparing Aortic Root Replacement

|  | Total Patients<br>(n=284) |
| --- | --- |
| **Type of Procedure** |  |
| Re-implantation (David's) | 270 (95.1%) |
| Re-modelling (Yacoub) | 14 (4.9%) |
| **Concomitant Surgery** |  |
| Any Cardiac | 113 (39.8%) |
| CABG | 39 (13.7%) |
| Mitral Valve | 36 (12.7%) |
| Arch Repair | 57 (20.4%) |
| **Leaflet Repair** | 200 (70.4%) |
| **Type of Leaflet Repair** |  |
| Central Plication | 128 (45.1%) |
| Peri-Commissural Plication | 23 (8.1%) |
| Gore-Tex to free edge | 40 (14.1%) |
| Wedge | 1 (0.4%) |

### Early Outcomes

The 30-day mortality was 1.8%, with 6 (2.1%) requiring mechanical support, 4 (1.4%) having a stroke, and 23 (8.1%) returning to theatre for bleeding. Other 30-day outcomes are as described in Table 3.

**Table 3.** 30-day outcomes of valve-sparing aortic surgery. *Abbreviations: ECMO = Extracorporeal Membrane Oxygenation; IABP = Intra-aortic Balloon Pump; RTT = Return to theatre*

| Outcome | Overall<br>(n=284) | Pre-2010<br>(n=100) | Post-2010<br>(n=184) | P value |
| --- | --- | --- | --- | --- |
| **30-day mortality** | 5 (1.8%) | 4 (4.0%) | 1 (0.5%) | 0.03 |
| **IABP/ECMO** | 6 (2.1%) | 5 (5.0%) | 1 (0.5%) | 0.01 |
| **Stroke** | 4 (1.4%) | 1 (1.0%) | 3 (1.6%) | 0.67 |
| **RTT for bleeding** | 23 (8.1%) | 10 (10.0%) | 13 (7.1%) | 0.39 |
| **Dialysis/Filtration** | 7 (2.5%) | 3 (3.0%) | 4 (2.2%) | 0.67 |
| **Tracheostomy** | 3 (1.1%) | 0 (0.0%) | 3 (1.6%) | 0.18 |

### Survival

Overall survival was 96.0% (95% CI 92.6-97.8%) at 5 years, 88.2% (95% CI 81.4-92.6%) at 10 years and 78.6% at 15 years (66.2-86.9%). Valve morphology (BAV vs. TAV) wasn’t a predictor of mortality (HR 1.08, 95% CI 1.01-1.09, p=0.91) in the multivariable cox regression model. The only predictors were older age at time of intervention (HR 1.05 per year increase, 95% CI 1.01-1.09, p<0.01) and ATAAD (HR 3.58, 95% CI 1.03-12.4, p<0.01), with male sex being protective (HR 0.30, 95% CI 0.13-0.08, p<0.01). Concomitant cardiac procedure and leaflet repair also did not predict mortality. Of all mortalities, 29% were cardiac, 13% secondary to a malignancy/cancer, 3% to a non-admission infection, with other causes as shown in Supplementary Figure 1.

### Re-intervention and Aortic Insufficiency

Overall rates freedom from re-intervention was 92.2% (95% CI 87.7-95.2%) at 5 years, 79.8% (95% CI 71.8-85.8%) at 10 years, and 74.1% (63.5-82.0%) at 15 years. The only predictors of re-intervention were concomitant leaflet repair at the time of VSARR (HR 8.13, 95%CI 1.07-61.7) and bicuspid aortopathy (HR 2.23, 95% CI 1.07-4.68) (Table 4). For BAV patients, this translates to a worse 5- (87.0% vs. 93.8%), 10- (62.3% vs. 84.7%) and 15- (43.6% vs. 82.8%) year freedom from re-intervention compared to TAV patients (Log-rank p<0.01) (Figure 2B). For patients who underwent concomitant leaflet repair at time of surgery, the 5-, 10- and 15-year freedom was lower at 89.9%, 75.0% and 67.4% compared to 98.7%, 94.6% and 94.6% respectively (Figure 2C). The freedom from aortic insufficiency was 89.1% (95% CI 83.5-92.9%), 84.9% (95% CI 77.8-89.9%) and 80.7% (71.0-87.4%) at 5-, 10- and 15-years respectively (Figure 2D). There were no statistically significant differences in rates of aortic insufficiency at 15 years when stratified by bicuspid valve morphology (Bicuspid 81.5% vs. Tricuspid 80.4%, P=0.60) and concomitant leaflet repair (Leaflet Repair 81.8% vs. No Leaflet Repair 78.7%, p=0.69). This was consisted with Cox-Regression analysis (Table 4), which demonstrated no statistically significant risk of VSARR for patients with BAV morphology (HR 1.76, 95% CI 0.76-4.09, Figure 2E) or who underwent concomitant leaflet repair (HR 1.70, 95% CI 0.58-5.01, Figure 2F).

**Table 4.**
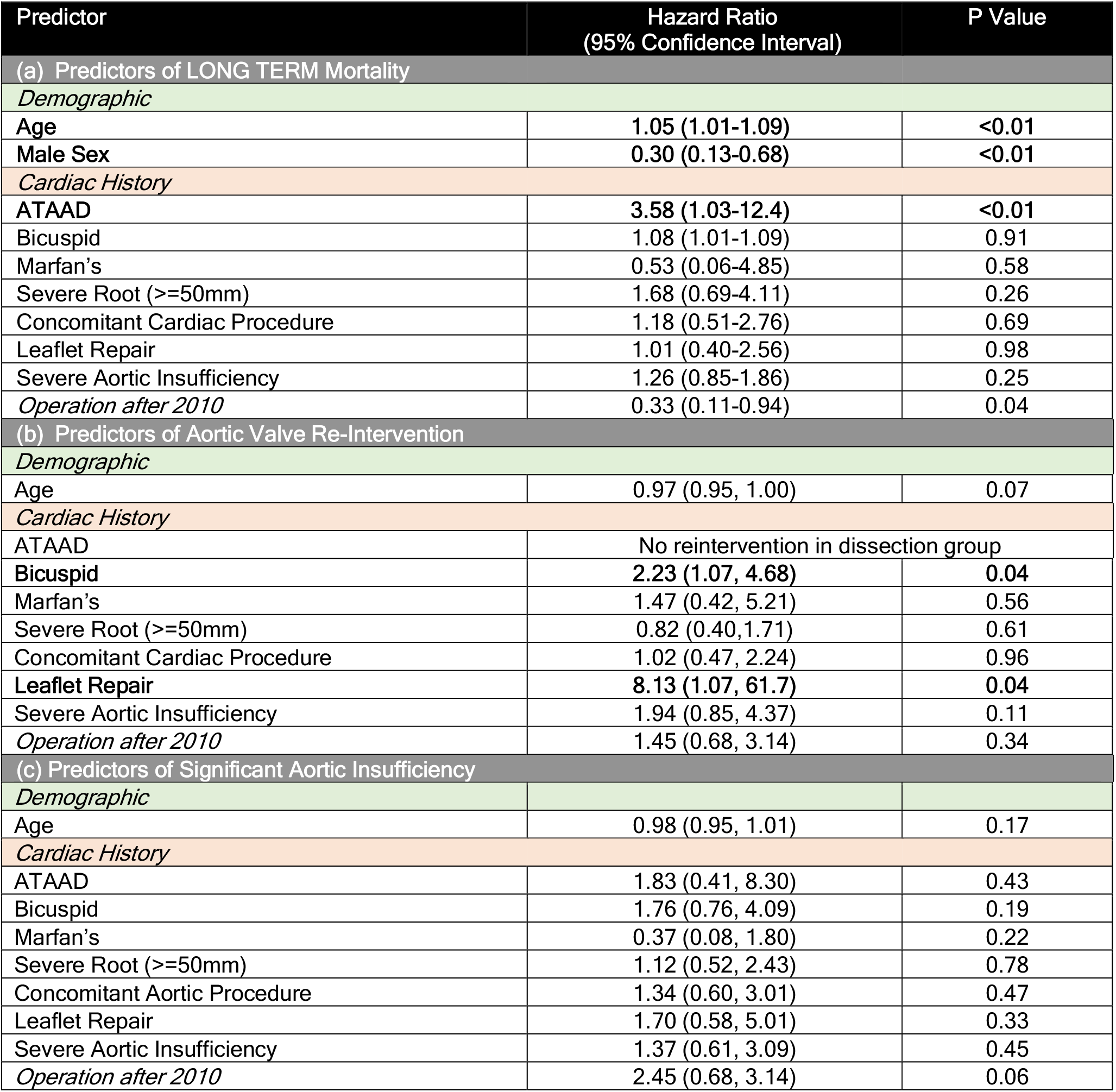
Multivariable Cox regression analysis for predictors of long term (up to 15 years) (a) Mortality, (b) Aortic Valve Re-intervention and (c) Significant Aortic Insufficiency.

**Figure 1.**
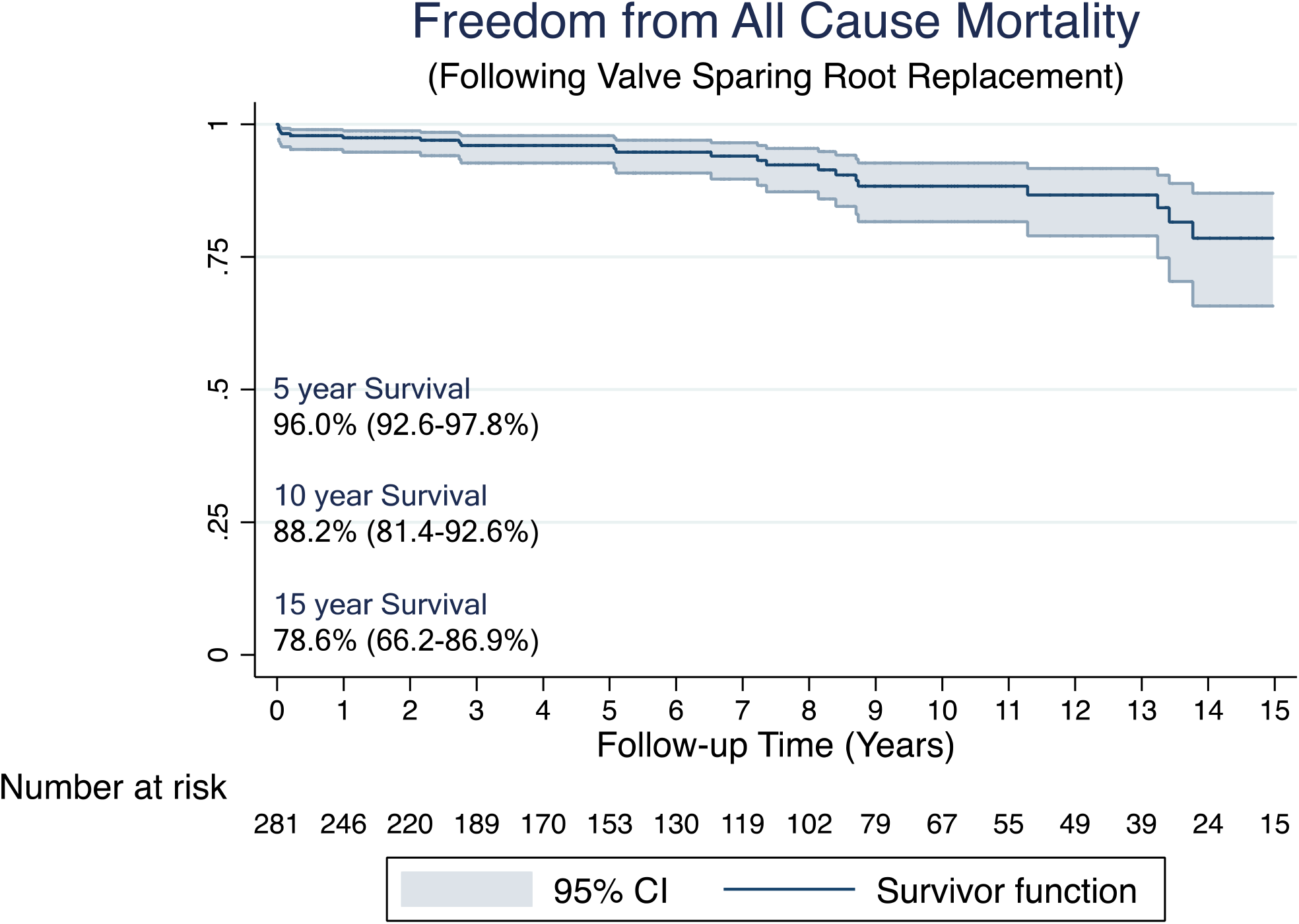
Freedom from all-cause mortality at up to 10 years

**Figure 2.**
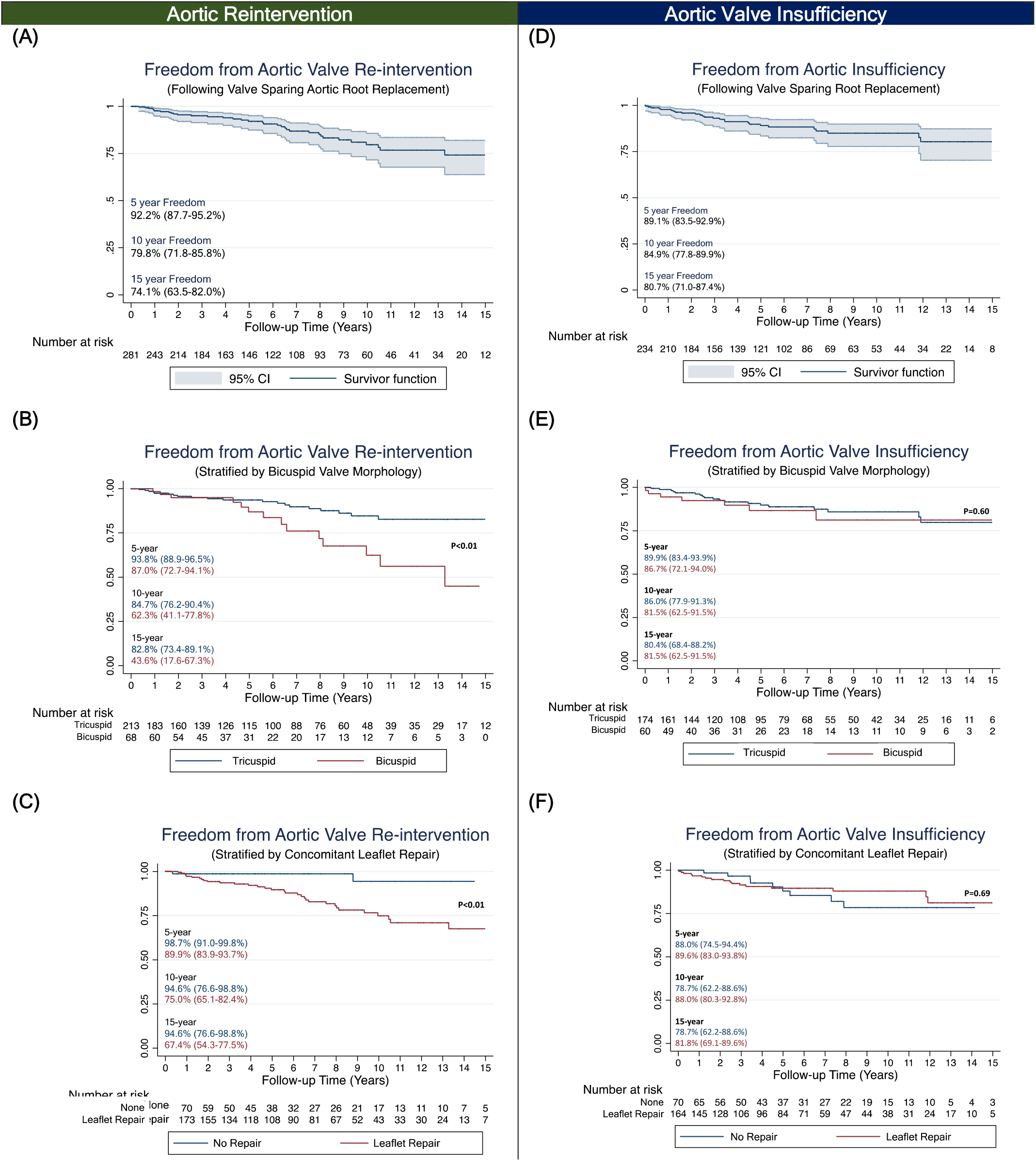
Freedoms from (A) Overall Aortic Re-intervention, stratified by (B) Bicuspid Valve Morphology and (C) concomitant leaflet repair. Freedoms from (D) Overall Aortic Valve Insufficiency, stratified by (E) Bicuspid Valve Morphology and (F) concomitant leaflet repair

### Era of Intervention

Patients who underwent intervention post-2010 had a significant reduction in peri-operative mortality (0.5% vs. 4.0%, p=0.03) and need for mechanical support (0.5% vs. 5.0%, p=0.01). In risk and time-adjusted analysis, post-2010 patients had one-third (HR 0.33, 95% CI 0.11-0.94, p=0.04) the risk of long-term mortality at any given year of follow up. There was no difference in the risk of long-term aortic insufficiency or re-intervention in post-2010 patients.

### Aortic Valve Re-intervention

The nature of re-interventions to the aortic valve and root is listed in Table 5. The most common was an isolated aortic valve replacement (n=23), followed by re-do aortic root replacement (n=6) for Infective endocarditis (n=3) or a false aneurysm (n=3), and then aortic valve replacement with ascending or arch repair (n=4) and a Ross procedure (n=1).

**Table 5.** Description of Aortic Re-interventions

| Aortic Re-intervention | Number of Interventions<br>(n=34) |
| --- | --- |
| Aortic Valve Replacement | 23 (67.6%) |
| Redo Aortic Root Replacement (including Aortic Valve) | 6 (17.6%) |
| Aortic Valve Replacement with Ascending/Arch Repair | 4 (11.8%) |
| Ross Procedure | 1 (2.9%) |

## Discussion

We have described the outcomes following valve sparing aortic root replacement (VSARR) in a series 284 consecutive patients. The procedure carries a low 30-day mortality of 1.8%. Furthermore, VSARR in the contemporary era (post-2010) carried one-third the hazard of death compared to those done in the preceding decade (HR 0.33 95% CI 0.11-0.94). It also demonstrated excellent long-term performance, with a 10-year survival of 88.2%. This compares favourably with survival after bioprosthetic and mechanical root replacement of 72 to 81% [5-7] at 10 years. Freedom from re-intervention and insufficiency were 79.8% and 84.9% respectively at 10 years. These are comparable to other published results [1-3, 5, 8-14], although few extend out to 15 years.

Predictors of the need for aortic re-intervention were BAV morphology and concomitant leaflet repair. In patients who do not need concomitant leaflet repair, our 15-year freedom from reoperation was 94.6%, supporting guidelines that VSARR should be the treatment of choice in patients who have a morphologically normal aortic valve not needing repair [3]. Concomitant repair significantly increases the hazard of long-term re-intervention (HR 8.13, 95% CI 1.07-61.7) and therefore needs to be undertaken judiciously on a case-by-case basis. For instance, we have learned from experience that valves with even early degrees of calcification had a higher medium term failure rate, while those with supple leaflets did well even with major leaflet repairs. Whilst bicuspid valve disease does not influence survival (HR 1.08, 95% CI 1.01-1.09, p=0.91), it places the patient at over double the hazard of re-intervention (HR 2.23, 95% CI 1.07-4.68), with significantly worse 15-year freedom from re-intervention (43.6% vs. 82.8%). Again, our experience suggests that bicuspid valves free of calcium or significant scarring of the raphe perform superiorly. In bicuspid patient with unfavourable leaflets, we would now electively perform the Ross operation in suitable patients.

Concerns over the negative impact of pre-operative severe root-dilation (>50mm) or severe aortic insufficiency [10] was not supported in our cohort. Our data also supports guidelines [2] suggesting VSARR can be safely offered for patients with genetic aortopathies, such as Marfan’s Disease. In our series Marfan’s disease did not impact long term survival or freedom from aortic re-intervention or insufficiency.

Given that operative risks display a learning curve (interventions in the second decade had a lower 30-day mortality of 0.5% vs. 4.0%, and carry one-third the hazard of long-term mortality over 15 years follow up), good case selection requires recognition of unfavourable morphological factors, and that many cases require concomitant major surgery (39.8% need other cardiac interventions, including 20.4% requiring arch repair), we believe that these procedures should be performed in a centre with experience in VSARR surgery.

The limitations of this study are inherent to those as a retrospective study. Further delineation of the mechanism of failure needing re-intervention, specifically stenosis or regurgitation, would better help delineate the natural long-term progression of this operation. We suspect that many patients with BAV failed in the medium term with mainly aortic stenosis as a result of accelerated progression of calcification in leaflets with pre-existing calcification or marked fibrosis. This is currently under investigation at our institute. The strengths of our series are complete 30-day outcome data, and complete follow up of all patients with echocardiographic verification of valve function status of no longer than 2 years of last visit, and providing one of the largest series of Valve Sparing Root replacement published to date.

## Data Availability

Can provide data at request

## APPENDIX

**Figure 1.**
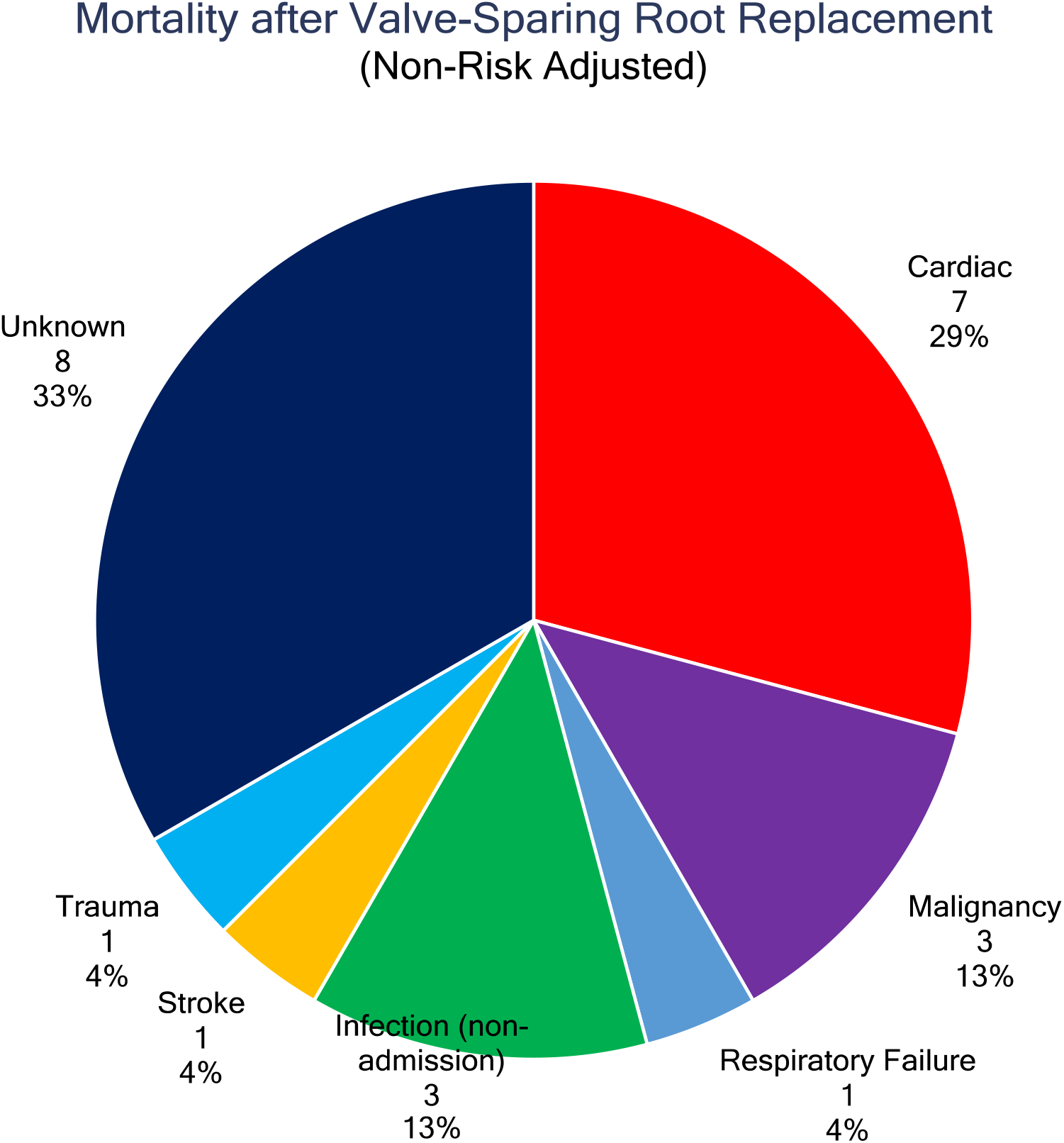
Causes of all mortality during this period

## Notes

### Competing Interest Statement

The authors have declared no competing interest.

### Funding Statement

Dr. Varun Sharma is a recipient of the National Heart Foundation of Australia PhD Scholarship

